# Acute Pulmonary Embolism in Critically Ill Patients with COVID-19

**DOI:** 10.1101/2020.05.22.20110270

**Authors:** Madhura Manjunath, Julio Miranda, Liana Fraenkel, Paul Mange Johansen, Blessing Phinney, Georgianne Valli-Harwood, Cynthia Callahan, Hafez Alsmaan, David Oelberg

## Abstract

Since the discovery of the novel coronavirus (SARS-Co-V-2) in December 2019, multiple characteristics have been reported, as our understanding of this new disease unfolds. One such association is its tendency to cause thromboembolic events, particularly venous thromboembolism^1,2^. In a four-week period during the initial spread of COVID-19 at a 300 bed community hospital in western Massachusetts, 23 patients who were PCR positive for SARS-CoV-2 RNA required treatment in either the intensive care unit (ICU) or intermediate/step-down unit (SDU). All patients were treated with standard DVT prophylaxis from the time of admission, except for two patients who were on full anticoagulation for chronic atrial fibrillation. Of the 23 patients, 7 (30%) were diagnosed with acute, clinically significant, pulmonary embolism (PE). Four of the 7 manifested evidence of acute cor pulmonale, one of whom succumbed as a direct consequence of a massive PE. Other markers were reviewed in the 7 patients to identify trends that could allow for early suspicion of PE in COVID-19 patients. Although D-dimer tended to rise during the hospitalization relative to the control group, the results were inconsistent, and there were no other meaningful distinguishing features between the groups at the time of admission.

## Methods

Starting in mid-March, we reviewed the number of patients who were critically ill from confirmed COVID-19 and required ICU or SDU level of care over a four-week period at Berkshire Medical Center in Pittsfield, MA. We compared the clinical features and laboratory test results of patients with and without PE. We were limited to descriptive analysis between the two groups given the small sample size. The study was approved by the institution’s IRB. Informed consent was waived based on criteria outlined in 45 CFR 46.104(d)(4).

## Results

Of the 23 COVID-19 positive, critically ill, patients hospitalized during a four-week period at our institution, 7 (30%) were diagnosed with concomitant acute PE during their inpatient stay. The two groups (PE+, n=7; PE-, n=16), had similar features on admission (Table 1). All patients in both groups were either former or never smokers. Respiratory and gastrointestinal symptoms were the most common complaints, but the mean resting heart rate was noted to be modestly higher in the PE group (104 vs. 88). Both groups showed lymphopenia. Pro-BNP was higher in the PE group at the time of admission, but the comparison was limited by incomplete data for the two groups. D-dimer trended upward in the PE group compared to the control group during the hospitalization (Figure 1). Other risk factors for PE, including BMI, recent surgeries, and travel history were similar across both groups.

**Table 1.**
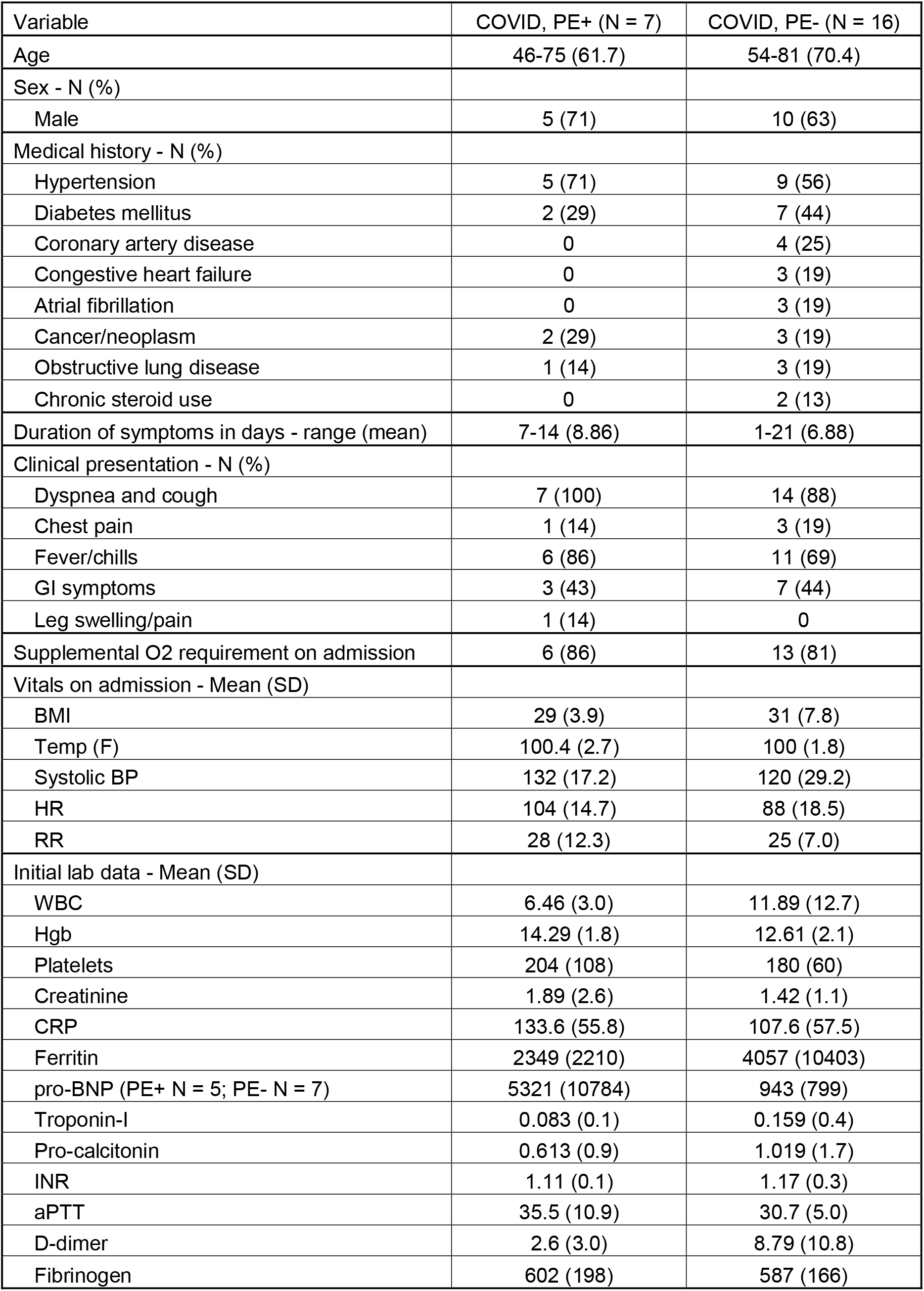
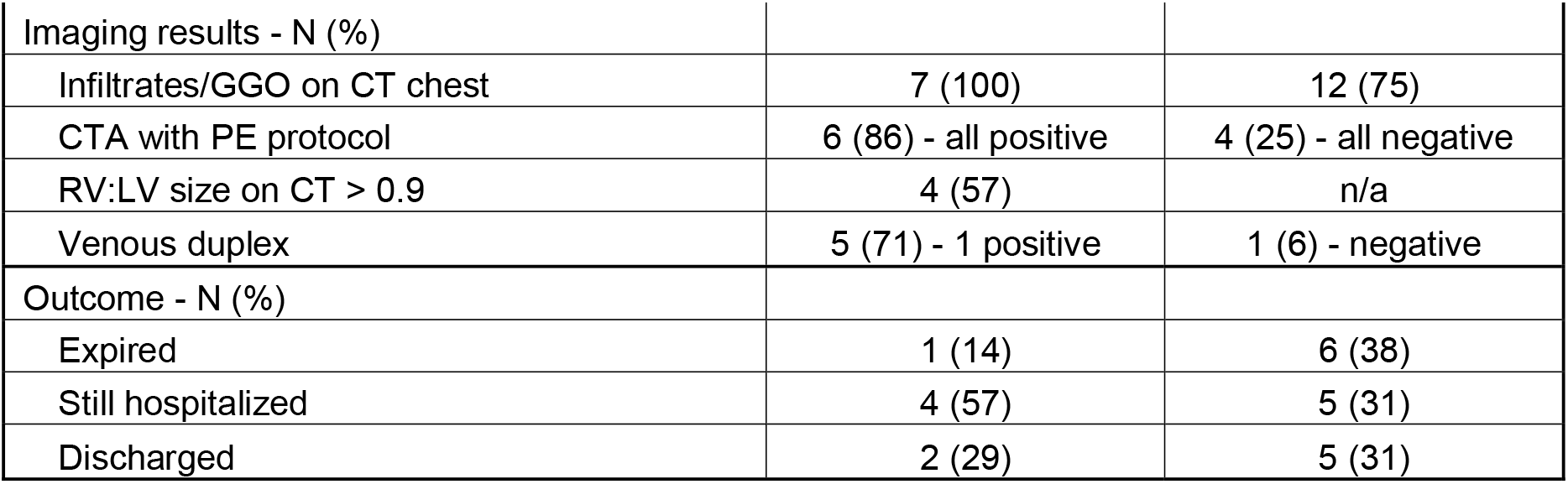

**Figure 1.**
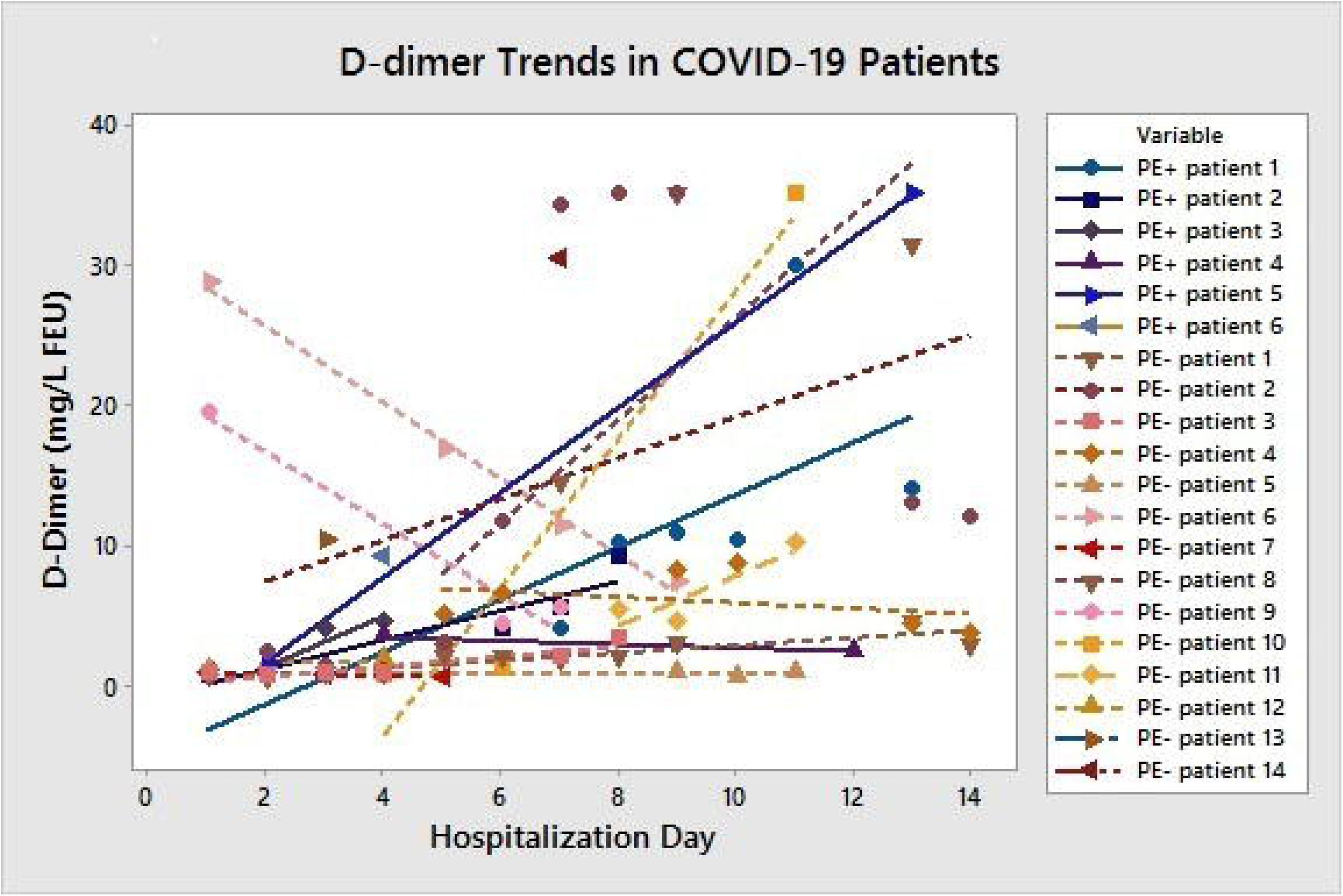
Of the 7 PE+ patients, one had no D-dimer measurements and one had a single D-dimer measurement (patient 6); of the 16 PE- patients, two had no D-dimer measurements and two had a single D-dimer measurement (patients 13 and 14). Solid linear regression lines represent PE+ patients (N=5) and dotted linear regression lines represent PE- patients (N=12). Decreasing linear regression lines only occurred in PE- patients (2 of 12 = 17%). Increasing linear regression lines occurred in 80% (4 of 5) of the PE+ patients and 33% (4 of 12) of the PE- patients. The remaining linear regression lines were essentially flat, occurring in 20% (1 of 5) of the PE+ patients and 50% (6 of 12) of the PE- patients.

Twenty one of the 23 patients were treated with DVT prophylaxis using either subcutaneous unfractionated heparin (5000 units BID, n=11) or low molecular weight heparin (40 mg daily, n=10) starting at the time of admission, while two patients were maintained on their home regimen of novel oral anticoagulants for atrial fibrillation (both in the PE-group). Of the seven patients who developed acute PE, the mean time to diagnosis was 9.4 days following hospitalization. The mean Wells score for the PE+ group was 5.7 and the PE-group 4.2, indicating a lack of discrimination with use of Wells criteria in these two groups. The diagnosis of acute PE was confirmed by CT pulmonary angiography in 6 patients and by transthoracic echocardiography in one patient, who suddenly became hypotensive on day five with severe right ventricular dysfunction and a positive McConnell sign. Despite treatment with thrombolytic therapy (100 mg intravenous tissue plasminogen activator) he expired three days later with multi-organ failure secondary to obstructive shock.

Of the 6 surviving PE patients, four were stratified as having minor (low risk) and two as having submassive (intermediate risk) pulmonary emboli. The mean pulmonary embolism severity index (PESI) score among our 7 PE patients was 119, indicative of high risk of mortality and morbidity (class IV, 4 to 11.4% 30-day mortality risk). Five received anticoagulation with either therapeutic doses of Heparin or Enoxaparin and were later converted to oral anticoagulation. One patient was found to have small, subsegmental PE on CTA, thought to be very low risk, and was maintained on prophylactic doses of subcutaneous Heparin (increased to 5000 units TID).

## Discussion

Our study adds to the growing recognition that patients with COVID-19 appear to be at high risk of developing PE. Since a search for PE was not conducted in all our patients admitted to the ICU, the 30% incidence of PE identified in this study may be an underestimate. The mechanism of hypercoagulability in these patients is not known, but the emergence of antiphospholipid antibodies and/or triggering of disseminated intravascular coagulation in critically ill patients have been suggested as possible mechanisms^3,4^. The latter is also supported by autopsy reports, which have shown the presence of frank hemorrhage along with small thrombi in sections of the lung parenchyma^5^.

In light of the growing incidence of acute PE in COVID-19 patients, despite standard DVT prophylaxis, intensified regimens of thromboprophylaxis may be indicated. D-dimer tended to rise during hospitalization in the PE group, which is consistent with studies correlating high D-dimer levels in patients at high-risk for PE^6^. However, the results were not consistent, so we could not conclude that initial or serial D-dimer assays could reliably predict who would subsequently develop PE and who would not.

The incidence of symptomatic PE in a general medical ICU is around 0.7-6%^7^, well below the 30% incidence reported in this series. Larger studies are needed to identify risk factors for PE and determine the level of anticoagulation required for specific subgroups. Meanwhile, clinicians should have a low threshold to suspect venous thromboembolism while treating patients with COVID-19 and consider routinely screening these patients and/or utilizing higher doses of thrombo-prophylactic regimens than the current standard for critically ill patients.

## Data Availability

Data summary is included in the manuscript as Table 1.

## Acknowledgements

MM, LF, and DO helped design the study based on the observations of GVH, HA, CC, JM, and DO in the ICU. Data collection was done through chart review by medical residents (see below) under the supervision of MM and BP. Analysis and interpretation of the data was aided by LF and PMJ. Drafting of the manuscript and the revisions were primarily done by MM and DO, with significant input from all other authors listed.

Medical residents involved with the chart review include Andrea Parra Corral, MD; Anoop Kumar, MD; Areej Khan, MD; Mintu John, MD; Muhammad Bilal, MD; Taroob Latef, MD; Xiaocao Xu, MD.

Mazen Ghani, MD with the Radiology department at Berkshire Medical Center reviewed the imaging to help us with readings and measurements.

**Abbreviation List**
COVID-19: coronavirus disease 2019
PCR: polymerase chain reaction
RNA: ribonucleic acid
ICU: intensive care unit
SDU: step down unit
DVT: deep venous thrombosis
PE: pulmonary embolism
IRB: institutional review board
CFR: code of federal regulations
BMI: body mass index
BID: twice daily
CT: computerized tomography
mg: milligram
PESI: pulmonary embolism severity index
CTA: computerized tomography, angiography
TID: three times daily
N: number
GI: gastrointestinal
O2: oxygen
SD: standard deviation
BP: blood pressure
HR: heart rate
RR: respiratory rate
WBC: white blood count
Hgb: hemoglobin
CRP: c-reactive protein
BNP: brain natriuretic peptide
INR: international normalized ratio
aPTT: activated partial thromboplastin time
GGO: ground glass opacities
RV: right ventricle
LV: left ventricle
n/a: not applicable
FEU: fibrinogen equivalent units

